# State Laws and Regulations Addressing Nurse-Initiated Protocols and Use of Nurse-Initiated Protocols in Emergency Departments: A Mixed-methods Cross-sectional Survey Study

**DOI:** 10.1101/2020.03.19.20038414

**Authors:** Jessica Castner, Lenore Boris

**Affiliations:** conducted this study as faculty with the University at Buffalo School of Nursing and is the President and Principal Investigator/Consultant for Castner Incorporated; Upstate Medical University College of Medicine

**Keywords:** emergency nursing, emergency department, scope of practice, protocol, nurse practice act

## Abstract

**Introduction:** State regulations may impede the use of nurse-initiated protocols to begin life-saving treatments when patients arrive to the emergency department. In crowding and small-scale disaster events, this could translate to life and death practice differences. Nevertheless, research demonstrates nurses do utilize nurse-initiated protocols despite legal prohibitions. The purpose of this study was to explore the relationship of the state regulatory environment as expressed in nurse practice acts and interpretive statements prohibiting the use of nurse-initiated protocols with hospital use of nurse-initiated protocols in emergency departments.

**Methods:** A mixed-methods approach was used with a cross-sectional nationwide survey. The independent variable categorized the location of the hospital in states that have a protocol prohibition. Outcomes included protocols for blood laboratory tests, x-rays, over the counter medication, and electrocardiograms. A second analysis was completed with New York State alone because this state has the strongest language prohibiting nurse-initiated protocols.

**Results:** 350 surveys from 48 states and the District of Columbia were received. A hospital was more likely to have policies supporting nurse-initiated protocols if they were not in a state with scope of practice prohibitions. Four qualitative categories emerged: advantages, approval, prohibition, and conditions under which protocols can be used. Prohibitive language was associated with less protocol use for emergency care.

**Conclusion:** State scope of practice inconsistencies create misalignment with emergency nurse education and training, which may impede timely care and contribute to inequalities and inefficiencies in emergency care. In addition, prohibitive language places practicing nurses responding to emergencies in crowded work environments at risk.

## Introduction

Comprising the largest sector of the professional health care workforce, registered nurses (RN) have immense potential to transform the health care system towards greater efficiency, cost-effectiveness, and higher quality care (Committee on the Robert Wood Johnson Foundation Initiative on the Future of Nursing, 2011). A registered nurse’s scope of practice is defined by the laws of each state. These generically termed nurse practice acts broadly outline the legal limits of registered nurses’ professional activities. Nurse practice acts vary by state. However, in many states, regulatory and policy barriers constrain nursing practice (Castner et al., 2013). These regulatory and policy barriers and inconsistencies often lead nurses to question whether certain activities are within their scope of practice. While regulatory and policy questions impact practical/vocational, registered and advanced practice nurses, the impact is especially poignant for registered nurses practicing in emergency departments (EDs). Here, we test the relationship between state RN scope of practice and RN informants’ perception of their employing hospitals use of nurse-initiated protocols in the emergency department. We further explore the RN’s qualitative comments about nurse-initiated protocols in the ED.

Emergency care is a crucial health care area where barriers to nursing practice scope and autonomy is particularly poignant, as demand for nursing care often outstrips resources and timeliness of care is essential to patient outcomes (Pines et al., 2011). In the ED, RNs are typically the first to assess a patient’s condition and are thus in ideal positions to initiate timely care, based on pre-established facility protocols (Castner et al., 2013; Douma, Drake, O’Dochartaigh, & Smith, 2016). This practice is often referred to as nurse-initiated protocols, complaint-specific protocols, advanced triage orders, medical directives, and standing triage protocols. Examples include administering nitroglycerin and obtaining an electrocardiogram (ECG) for chest pain, initiating standard laboratory testing on the blood samples of patients with signs and symptoms of an acute stroke, or ordering simple x-rays and administering over the counter pain relievers to a suspected wrist fracture (Bruce, Maiden, Fedullo, & Kim, 2015; Considine, Shaban, Curtis, & Fry, 2019; Ho, Chau, Chan, & Yau, 2018).

Frequently in the ED setting, the prescribing provider is unavailable to see the patient in a timely manner due to other, high priority tasks. Rapid initiation of interventions and diagnostic testing are essential to patient outcomes, and the emergency nurse has adequate preparation and training to prudently initiate treatment and tests (Anderson et al., 2007; Retezar, Bessman, Ding, Zeger, & McCarthy, 2011; Rivers et al., 2001; Rowe et al., 2011). In these instances, RNs may initiate treatment and diagnostic testing based on patients’ presentations only when both the state scope of practice regulation and hospital policy allow nurse-initiated protocols. However, there are many inconsistencies among states in how the RN scope of practice is regulated that prohibit RNs from applying these protocols to a patient without a patient-specific written or verbal prescription (Castner et al., 2013).

## Background

The delivery of health care is highly regulated, with hospital policy shaped by federal and state legislation and regulations. State regulation of nursing is a product of laws, regulations, and interpretive rules and statements of state nursing practice governing boards. While government and non-government agencies, such as Centers for Medicare and Medicaid Services (CMS) and the Joint Commission do not directly regulate nursing practice, a history of fining and penalizing facilities for nurse-initiated protocols effectively limited protocol use despite the fact that such protocols enhance efficiency and timeliness of care in the ED. In response to pressure from emergency medicine and nursing groups, CMS issued a letter clarifying prior policy directives and asserting the conditions under which nurse-initiated protocols can be used (CMS, 2008; Institute of Medicine [IOM], 2006).

The language of state scope of practice acts was analyzed in a previous study (Castner et al., 2013). Three major themes were identified that applied to nurse-initiated protocols in the ED setting: cautiously within scope, intentionally vague/silent, and outside scope of practice (7 states of CT, KS, NY, OK, IL, LA, NJ). States, such as Nevada and South Dakota, where the scope of practice language described nurse-initiated protocols as being cautiously within scope indicated that nurse-initiated protocols were appropriate for practice when justified by national and specialty standards, scientific literature, and accreditation standards. For the RN to initiate a set of treatment and diagnostic prescriptions to a patient before a relationship with a prescribing provider is initiated, there must be a guideline or protocol in place that is approved and delegated to the RN by the institution’s medical and nursing leadership (Lawrence, Scaletta, & Mason, 2007). Regulatory language that appears intentionally vague or silent, such as in Indiana or Pennsylvania, often provided general decision-making algorithms, without mentioning specific tasks, which emphasize the need for prudent training and competency while allowing nursing practice to expand and evolve (Castner et al., 2013). Finally, in the United States, seven states in which the nurse scope of practice act or interpretive statements specifically prohibited the RN from applying nurse-initiated protocols, except in very limited public health circumstances (Castner et al., 2013). These states include New York and Oklahoma.

New York State documents had the clearest and strongest explicit prohibition of nurse-initiated protocols, with specific legislation and interpretive statements (Office of the Professions of New York State Education Department [OP], 2009). The only nurse-initiated protocols allowed in New York included the administration of immunization, anaphylactic agents, purified protein derivative tests, and human immunodeficiency virus tests. However, these circumstances apply only when the prescribing provider had no treatment relationship with the patient. What constitutes a treatment relationship is open to interpretation. A broad interpretation is that an attending ED physician’s treatment relationship begins when the patient arrives, even if the ED physician is not yet aware of the patient, rather than a more limited view, the relationship begins when the ED physician actually starts treating the patient. When there are multiple ED prescribing providers on site at the same time, a broad interpretation becomes more complex.

Several hospital quality metrics rely on ED efficiency beginning as soon as the patient arrives (Leveille et al., 2020; McClelland, Jones, Siegel, & Pines, 2012). The patient with an acute heart attack’s time from door to treatment, the patient with stroke symptoms time to medication, or the septic patient’s time to antibiotics all impact patient outcomes and hospital quality ratings. Because the RN is often in the best position to initiate urgent care in the ED, the timeliness of this lifesaving care is facilitated by nurse-initiated protocols. Inefficiencies can lead to ED crowding, which is linked to higher mortality, uncontrolled pain, and patient dissatisfaction (Bernstein et al., 2009; Collis, 2010; Sepahvand, Gholami, Hossenabadi, & Beiranvand, 2019; Sun et al., 2013). The health care and public health safety net, the ED setting is often crowded with responsibilities beyond capacity in both every day and small scale disaster response operations, where decisions can result in the difference between life and death.

Studies on nurse-initiated protocols demonstrate these protocols enable process and outcome improvements by reducing length of stay and improving patient outcomes (Considine et al., 2019; McArthur III & Thomas, 1995; Retezar et al., 2011; Rivers et al., 2001; Rowe et al., 2011; Seaberg & MacLeod, 1998). However, these relationships are not consistent for all types of patients. For example, RNs ordered more diagnostic x-rays and delayed the treatment for patients who present with abdominal pain, compared to their physician counterparts.

In summary, evidence generally supports the use of nurse-initiated protocols to enhance ED efficiency, with specific exceptions. With variations by state resulting in an inconsistent regulatory environment, the Emergency Nurses Association (the specialty organization for emergency nurses) supports the use of nurse-initiated protocols to improve patient flow through the ED (Emergency Nurses Association [ENA], 2014). Thus, states that prohibit nurse-initiated protocols create inconsistency with providing the best evidence practice, thereby placing both the providing RN and hospital in a morally and ethically ambiguous situation when a prescribing provider is not immediately available. It is unclear how the inconsistencies in state scope of practice regulation influences specific hospitals’ decision to allow the use of nurse-initiated protocols. The purpose of this study was to explore the relationship of the state regulatory environment as expressed in nurse practice acts and interpretive statements with hospital use of nurse-initiated protocols in emergency departments.

## Methods

### Design

A mixed-methods approach was used with a cross-sectional United States nationwide survey. The protocol (#547766-1) was approved by the Social and Behavioral Sciences Institutional Review Board at the University at Buffalo on January 15, 2014.

### Instrument

The survey was developed for the purposes of this study. Creating an original questionnaire was justified as no published or tested instrument was available to address the objective of the project. The survey focused on the hospital’s characteristics and policies because participants were considered informants of their hospital environments. The survey was self-administered electronically from January to June, 2014 through an Internet link using Vovici software. The survey included an open-ended field for comments after a yes/no binary response option to a question if the RN could initiate activities based on protocols established by the hospital.

### Participants

We used a combination of convenience and snowball recruitment methods. A multi-method approach was used to recruit participants using email invitations, an internet link on the Emergency Nurses Association Website, and social media. An email invitation was sent to the Emergency Nurses Association’s State-level leadership and to a purchased list of 1,243 nurse executives. The email invitation included a request to forward the email/link to those with clinical or management responsibilities for the ED in their respective organizations. For the purchased list, a reminder email was sent one week after the initial email. A third and fourth reminder email was sent to 980 nurse executives in the 24 states with the lowest numbers of responses. In addition, the survey link was posted on the Emergency Nurses Associations’ webpage for External Research Opportunities, and through social media (Facebook and Twitter) at the lead author’s university.

### Variables

The outcome (dependent) variables are the presence of the following nurse-initiated protocols: blood laboratory tests, x-rays, over-the-counter medications, and electrocardiograms. The independent variable was an indicator representing states where nurse-initiated protocols were prohibited by the scope of practice documents, one for New York State alone and one for all seven states with language that prohibits protocols (7 states of CT, KS, NY, OK, IL, LA, NJ). Model covariates included the respondent’s reports for number of certified hospital beds and the following categorical hospital characteristics: academic/teaching vs. non-academic/teaching, for profit vs. not-for-profit, specialty vs. general, and urban vs. non-urban.

### Sample Size Justification

According to Bell and colleagues, a survey sample size of 321 has sufficient power to generalize to the emergency nursing specialty in the United States (Bell, Dake, Price, Jordan, & Rega, 2014). We continued recruitment until reaching this sample size.

### Analysis

The unit of analysis was the individual informant. Information was not gathered about the informant personally, but about their employing hospital. Data were screened to determine if more than one informant was reporting the same hospital characteristics. Descriptive statistics were conducted on all variables. We fitted logistic regression models to evaluate the relationship between the state scope of practice prohibition and the hospital presence of nurse-initiated protocols. Data were analyzed with and without outliers. Qualitative comments were grouped using a thematic analysis.

## Results

A total of 350 surveys were received. One was excluded as the responses were an exact duplicate of a previously submitted survey (submitted only 2 minutes prior). Six surveys reported identical hospital characteristics to one other survey. Because the respondents were not asked to name their affiliated hospital, it is unclear if there were two informants reporting on the same hospital or two informants in hospitals with identical characteristics. Thus, these six surveys remained in the analysis. We removed outliers with more than 800 inpatient beds, which increased the model fit (using the -2LL and Nagelkerke R squared). With the exception of over-the-counter medications, removal of outliers did not change the significance of the overall model, nor the significance of the explanatory variables. 90 participants left qualitative comments.

### Quantitative Results

A total of 350 surveys were returned from 48 states and the District of Columbia (no responses were received from informants in hospitals from Delaware and New Mexico). Of the 349 responses used in the analysis, 63% were staff ED nurses (n=221), and 26% were hospital managers or administrators with ED responsibilities (n=92). The remaining 11% of informants included educators, physicians, nurse practitioners, affiliated nursing faculty, staff nurses in non-ED departments, or administrators with no ED responsibilities.

Table 1 lists the characteristics of the hospitals about which the hospitals were reporting. The majority (87.1%) were general, non-academic/teaching, not-for-profit hospitals. Nearly one-fourth (22.3%) of the informants were from states where the nurse practice act and interpretive statements prohibit nurse-initiated protocols before the prescribing provider applies the protocol to a specific patient. The majority of the hospitals represented had protocols for nurses to initiate blood laboratory studies, x-rays, over-the-counter medications, and ECGs before the prescribing provider interacted with the specific patient. Less than half also reported nurse-initiated protocols for more complex radiology studies or prescription medications.

**Table 1.** Characteristics of Hospitals (N=349)

| Characteristic | n | % |
| --- | --- | --- |
| Service Type |  |  |
| General | 304 | 87.1 |
| Specialty | 42 | 12.0 |
| Academic/Teaching | 143 | 41.0 |
| Non-Academic/Teaching | 199 | 57.0 |
| Profit Status |  |  |
| For Profit | 76 | 21.8 |
| Non- Profit | 264 | 75.6 |
| General Geographic |  |  |
| Urban | 150 | 43.0 |
| Suburban | 96 | 27.5 |
| Rural | 99 | 28.4 |
| Nurses Allowed to Initiate Protocols With |  |  |
| Laboratory Studies | 280 | 80.2 |
| X-Rays | 274 | 78.5 |
| Other Radiology | 60 | 17.2 |
| Over-the-counter Medications | 246 | 70.5 |
| Prescription Medications | 170 | 48.7 |
| Electrocardiogram | 286 | 81.9 |
| Located in State Where Nurse Practice Act Prohibits Protocols | 78 | 22.3 |
| Located in New York State | 53 | 15.2 |

The results of the logistic regression analysis are presented in Tables 2 and 3. New York State was compared to all other states, those with and without prohibitive language. All of the models were statistically significant and explained 9-39% of the variance in hospitals for nurse-initiated protocols. Regarding laboratory orders, hospitals in the 47 represented states other than New York demonstrated 5.7 times higher odds of having nurse-initiated protocols compared to New York State. Hospitals outside of the seven states with prohibitive scope of practice language demonstrated 3.6 times higher odds of having nurse-initiated protocols than the seven states with prohibitive scope of language practice. No other covariates predicted the differences in hospital policies. In regard to nurse-initiated protocols to prescribe x-rays, hospitals outside of New York State had 12.1 times higher odds of having protocols, while those outside the seven prohibitive states had 5.7 times higher odds of having protocols. Not-for-profit hospital status predicted a 2.6 times greater odds of nurse-initiated protocols, when comparing outside to inside New York State hospitals. For over-the-counter medication administration, both hospitals outside New York State (4.6 times) and those outside of the seven prohibited states (3.0 times) had higher odds of having nurse-initiated protocols than New York State and non-prohibitive states, respectively. Finally, for ECGs, hospitals outside of New York State (5.6 times) and the seven prohibited states (3.3 times) demonstrated higher odds having nurse-initiated protocols than the comparison states.

**Table 2.**
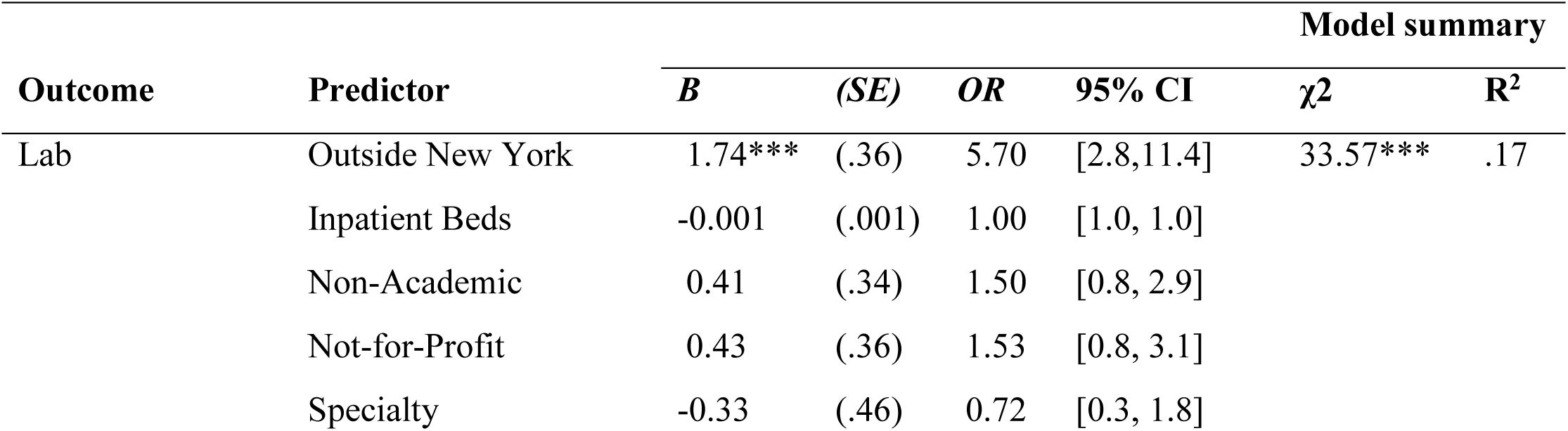
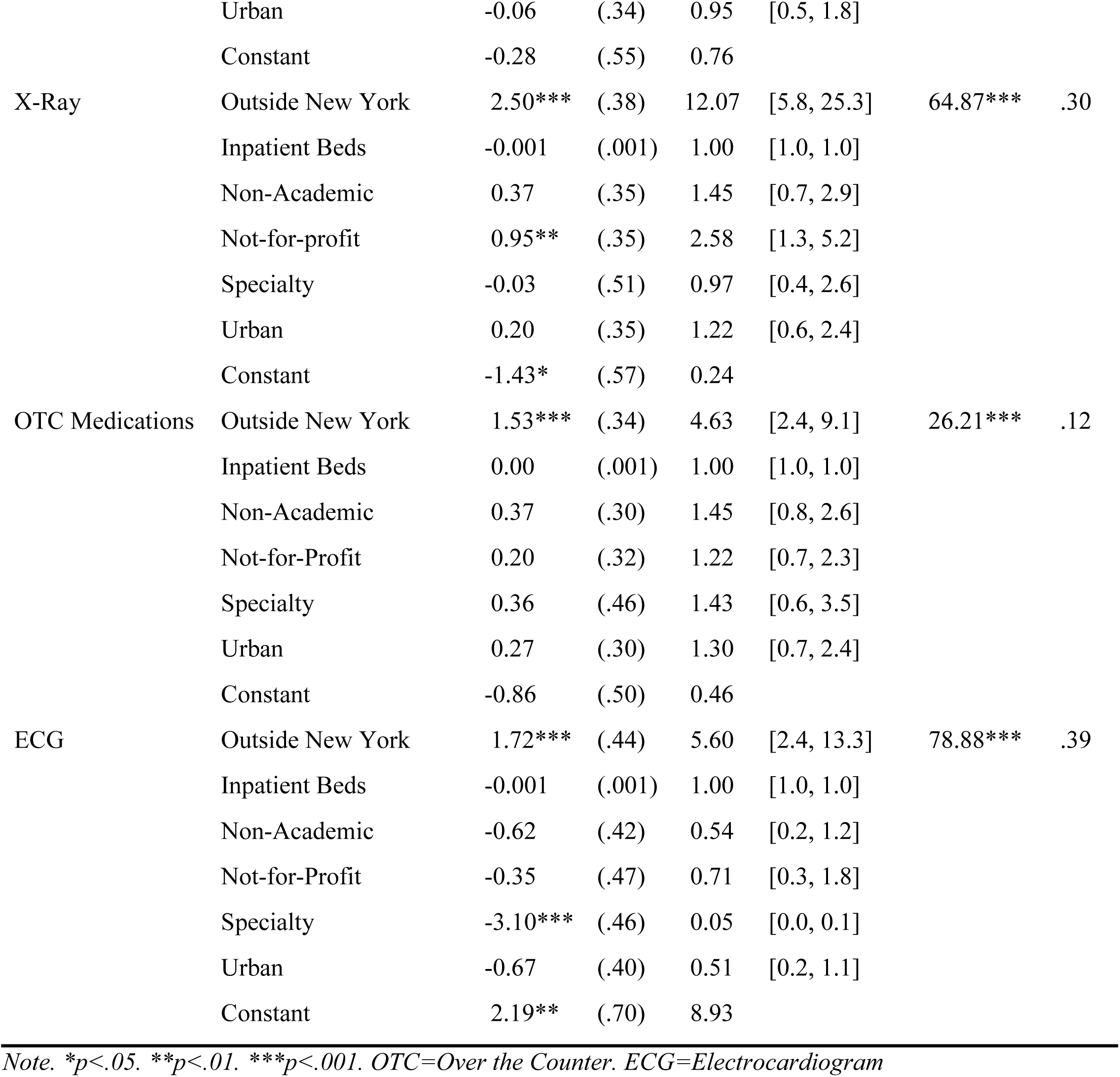
Logistic Regression Models for Inside vs. Outside New York State Hospitals and Nurse-Initiated Protocols

**Table 3.** Logistic Regression Models for Protocol Allowed vs. Prohibited State Hospitals and Nurse-Initiated Protocols

| Outcome | Predictor | Model summary |  |  |  |  |  |
| --- | --- | --- | --- | --- | --- | --- | --- |
| | | *B* | (*SE*) | *OR* | 95% CI | $\chi^2$ | $R^2$ |
| Lab | Protocols Allowed | 1.26*** | (.32) | 3.58 | [1.9, 6.7] | 25.52*** | .13 |
|  | Inpatient Beds | 0.00 | (.001) | 1.00 | [1.0, 1.0] |  |  |
|  | Non-Academic | 0.42 | (.34) | 1.53 | [0.8, 3.0] |  |  |
|  | Not-for-Profit | 0.49 | (.35) | 1.64 | [0.8, 3.2] |  |  |
|  | Specialty | -0.33 | (.45) | 0.72 | [0.3, 1.8] |  |  |
|  | Urban | -0.26 | (.33) | 0.77 | [0.4, 1.5] |  |  |
|  | Constant | 0.16 | (.51) | 1.17 |  |  |  |
| X-Ray | Protocols Allowed | 1.74*** | (.32) | 5.69 | [3.1, 10.6] | 47.24*** | ..22 |
|  | Inpatient Beds | -0.001 | (.001) | 1.00 | [1.0, 1.0] |  |  |
|  | Non-Academic | 0.40 | (.34) | 1.50 | [0.8, 2.9] |  |  |
|  | Not-for-profit | 0.99** | (.34) | 2.68 | [1.4, 5.2] |  |  |
|  | Specialty | -0.04 | (.48) | 0.96 | [0.4, 2.5] |  |  |
|  | Urban | -0.12 | (.33) | 0.89 | [0.5, 1.7] |  |  |
|  | Constant | -0.69 | (.51) | 0.50 |  |  |  |
| OTC Medications | Protocols Allowed | 1.10*** | (.29) | 2.99 | [1.7, 5.3] | 19.6** | .09 |
|  | Inpatient Beds | 0.00 | (.001) | 1.00 | [1.0, 1.0] |  |  |
|  | Non-Academic | 0.38 | (.30) | 1.47 | [0.8, 2.6] |  |  |
|  | Not-for-Profit | 0.25 | (.31) | 1.28 | [0.7, 2.4] |  |  |
|  | Specialty | 0.34 | (.45) | 1.41 | [0.6, 3.4] |  |  |
|  | Urban | 0.10 | (.29) | 1.10 | [0.6, 2.0] |  |  |
|  | Constant | -0.42 | (.47) | 0.65 |  |  |  |
| ECG | Protocols Allowed | 1.20** | (.41) | 3.32 | [1.5, 7.4] | 72.53*** | .36 |
|  | Inpatient Beds | -0.001 | (.001) | 1.00 | [1.0, 1.0] |  |  |
|  | Non-Academic | -0.59 | (.42) | 0.55 | [0.2, 1.3] |  |  |
|  | Not-for-Profit | -0.22 | (.46) | 0.80 | [0.3, 2.0] |  |  |
|  | Specialty | -3.01*** | (.46) | 0.05 | [0.0, 0.1] |  |  |
|  | Urban | -0.84 | (.39) | 0.43 | [0.2, 0.9] |  |  |
|  | Constant | 2.57*** | (.68) | 13.11 |  |  |  |

*Note. \*p<.05. \*\*p<.01. \*\*\*p<.001. OTC=Over the Counter. ECG=Electrocardiogram.*

### Qualitative Results and Interpretation

All survey participants were provided fields for additional comments to the question asking if the RN could initiate activities based on protocols established by the hospital. The comments reflect survey participants’ thoughts and experience with nurse-initiated protocols without differentiating whether the nurse practices in a state that supports or prohibits use of nurse-initiated protocols. The comments fall into four broad categories that include *advantages, approval, prohibition*, and *conditions* under which protocols can be used.

### Advantages

Some advantages to nurse initiated protocols are summarized in the following comment “*nurses are encouraged to start protocols to help decrease wait time, length of stay, and [improve] patient satisfaction*”. One nurse, whose prior experience at a facility that gave nurses the autonomy to initiate protocols, write that use of nurse initiated protocols “*proved to increase the efficacy of treatment and patient flow*”. Perceived benefits to patients were commented on but vague. “*The ability of an RN to initiate triage protocols benefits the patient and their stay in the ED*” is one such comment with no indication of the types of benefits that accrued to the patient. Another nurse noted a hospital could not possibly achieve its goal of ten minutes from door to EKG time if the nurse had to wait for a prescription then added a qualifier “*not sure if it’s legal but we do it*”. Another nurse verified the reduction in patient wait time “*there have been limited protocols in the past but we have come quite a ways toward speeding up the process to cut down the time patient is in the ED*”.

### Approval

Comments spoke to the involvement of federal, state and/or facility involvement in the decision to use protocols or the need to approve protocols. At the federal level, a CMS visit to one hospital limited protocols such that the survey respondent noted “*now nursing is only allowed to order 5 OTC [over the counter] medications, a few x-rays and urine dip*”. Prior to the visit there were many more “*robust protocols*” in use. At the state level, Texas has standing delegation orders “*intended for pre-provider care*” The “*protocol is only used after an order to initiate a protocol is given by a provider*”. Similarly, nurses are not able to initiate protocols in an ED without a verbal conversation with a provider as required by an unspecified state’s Department of Education. Another comment mentioned “*our state Board of Nursing is silent on this* [the topic of nurse initiated protocols].” Silence by state boards of nursing on the issue is common (Castner et al., 2013).

Details of the institutional approval process or nurse initiated protocols were lacking as illustrated by the comment “*specifically allowed procedures and medications are included in written protocols that have been reviewed and approved*”. Approval of standardized protocols within the facility prior to implementation seems to be the sole purview of physicians. No mention was made of a process for developing nurse-initiated protocols that required approval by the Chief Nurse Executive or Chief Nursing Officer or involvement of committees that include nurses. Comments such as “*nurse initiated protocols went before the physician group and they signed off*” or “*advanced patient care protocols*…*are covered by acceptance by medical practice/review*” acknowledged protocols as physician prescriptions that required approval from physicians prior to implementation. Physician approval can be problematic even within the same organization as one nurse noted “*we have other protocols that have been approved by med staff but our ED leadership has not decided if they want to use them yet*”. A variation occurs post adoption “*these orders should be used by triage nurse but we still have some issues with individual practitioners not on board while others are upset when these are not initiated so we are struggling with consistency*”. Implementation can require the buy-in from medical administration, as evidenced in the remark “*we used to be able to [initiate protocols] but the medical director abolished the standard*”.

One comment noted “*protocols orders are policy driven*” The ready availability of a physician or an advanced practice provider mitigated the need for nurse protocols, as one nurse stated, “*we use a physician in triage 8 hours of the day, 10 am -6:00pm. This reduces the need for nurse initiated protocols*”. Another nurse described their situation with more detail “*we no longer are able to place orders per protocol without a provider order. We now have provider at triage. Where a provider, md [medical doctor], np [nurse practitioner] or pa [physician assistant], is stationed with the triage nurse and see the patient directly after triage. That provider places basic orders based on complaint, however does not follow the patient throughout their ED stay*”. These comments are in stark contrast to the nurse who lamented “*I wish there were more!*”.

### Prohibition

Some nurses commented about their hospital’s absolute prohibition against nurse-initiated protocols, “*we have no standing protocols*”, “*none*”, and “*unable to initiate any protocols*”. One comment reflected a move away from nurse-initiated protocols “*we no longer do any nurse driven protocols*” with no explanation for imposition of greater restrictions on nurse-initiated protocols.

Comments addressed initiating actions despite an absence of standing orders and prior to the physician assessing the patient and placing treatment orders. One nurse explained the situation thusly “*we have a very small ER and if the RN would like something before the MD sees the patient typically they just ask and the MD says ‘ok’*”. In some cases, the situation may appear to be that of an experienced nurse seeking a verbal prescription order from a physician. Verbal prescription orders hold potential for misunderstanding and thus are often discouraged. Other comments suggested the circumstances the ED nurses find themselves in describe a more complex situation. Another nurse provided more detail stating “*based on your relationship with the physician you can order more knowing their treatment practices and their comfort level with you as a competent nurse*”. The nurse clearly relayed some discomfort with reliance on the physician’s opinion of the nurse and need for the physician to be willing to backup any actions taken independently by the nurse for the comment went on to state “*misuse of this could lead to scope of practice issues so I do this in moderation”*. The lack of a relationship between the physician and patient, the alignment of the physician’s assessment with the nurse’s assessment, the willingness of the physician to allow the nurse to initiate treatment protocols, the availability of protocols, the direction or lack of direction on the issue of nurse–initiated protocols in state laws and regulation, and other factors confound a nurses ability to function in a situation where nurse-initiated protocols appear to be unofficially sanctioned.

Nurses also spoke about the pressing need in some situations to respond to the patient’s precarious medical condition quickly when a physician was not available. One comment summarized this sort of effort “*we have ‘ED standing orders’ or protocols in which we can order ourselves if a patient has not been assessed by a provider within an hour. Most nurses will add orders sooner than that and most providers appreciate the diagnostic tests already being completed and resulted after they just assessed them*”. Despite the absence of any written standing prescription or treatment protocols that get implemented with a physician’s verbal or written prescription, one nurse offered “*though not covered by a policy for protocols, for critically ill patients, nurses initiate labs, ECG, etc. until MD is available to respond to bedside”*.

Instead of “protocols”, several comments referenced “*order sets*” such as “*our ER has standing order sets based on chief complaint*” or “*there are order sets based on chief complaint, patient condition*”. Order sets appear to be another name for a prescribed set of actions that can be taken in response to specific patient complaints. If there are differences within an institution or between institutions in the defining characteristics (purpose, development and approval process, implementation, etc.) of protocols or standing protocols or order sets these were not addressed in the nurses’ comments.

In institutions that have approved protocols, considerable range in the number and intent for the protocols was discussed in the comments provided. Strict limitations in the number of protocols exist in some facilities “*the only protocol currently initiated in triage is for ECG if the patient has a presenting complaint of chest pain, palpitations, weakness, dizziness, non-traumatic arm/jaw pain or syncope/near syncope*”. In contrast, some hospitals have 25 or more nurse-initiated protocols as mentioned “*we have a list of about 40-50 protocols order sets that we can initiate from*”.

### Conditions

The purpose of protocols included treatment or testing for life threatening conditions, expediting tests for frequently seen conditions or relief of pain or some other symptom. For example “*RN may call RT [respiratory therapy] to begin respiratory treatment for acute respiratory distress, RN may also activate relevant ‘alerts’ and related protocol: i*.*e. stroke alert will generate order for neurology response and CT scan*”. Lab work and x-rays were two commonly mentioned tests the nurse could order. In some instances, lab work could be drawn but would “*be sent to [to laboratory for processing] when order obtained*”. Treatment of symptoms might include insertion of a catheter for urinary retention, oxygen therapy for respiratory problems or obtaining intravenous access for chest pain.

The ability for the RN to give medications to patients varied widely. One nurse noted “*the authority with meds is limited” citing a “more collaborative approach*”. More typically several types of pain medication including aspirin, Tylenol, Motrin, Morphine fentanyl intravenously were mentioned with one person stating “*we give a lot of narcotic pain meds per triage protocol*”. Other types of medications were given for nausea or as part of a protocol addressing diabetic or respiratory conditions.

From the comments it appears the use of some kind of protocols or prescription sets, formal or informal were in use in most of the facilities. Their use, however, did not grant ED registered nurses unrestricted ability to initiate the protocols. The most frequently listed limitation to the initiation of protocols by the nurse was the need for, at a minimum, a verbal prescription order. There were exceptions “*we have protocols but some require at least a verbal order to initiate them. Some we can start on our own*”. More typically “*we cannot initiate any orders without a physician approval except HIV testing if the patient requests it*” or “*we cannot as RN’s give medications or order any tests unless we are given verbal or written orders*”.

Some of the actions nurses-initiated without physician prescription orders seem relatively benign. Over the counter and non-narcotic pain relievers, ordering laboratory tests, sending a patient for an EKG and so on, nevertheless, are commonly recognized as tools used by physicians to accurately diagnose and appropriately medically treat a patient’s condition. Thus historically, in institutions, even seemingly benign or non-invasive actions could not be initiated by a nurse absent a physician’s prescription to the individual patient. Other factors, for example, the cost of procedures that may or may not be needed after a full patient assessment by the physician, the training received by nurses or the availability of adequate hospital resources may influence the decision to utilize nurse-initiated protocols.

## Discussion

The purpose of this study is to explore the relationship of the state regulatory environment, through the nurse practice act and interpretive statements, with hospital use of nurse-initiated protocols in the ED. In this study, we received surveys from 350 health care provider informants from across the United States about nurse-initiated protocols in their employing hospital environment.

We demonstrated a significant relationship between prohibitive language in the state nurse practice documents and the absence of hospital policies and protocols for nurse-initiated care in the ED, before a relationship is established with the provider. This has important implications for how quickly care can be provided in life-threatening situations. The associations were most profound for New York State, where explicit legislation limits RN autonomy and a facility’s ability to delegate protocols to the RN (OP, 2009). In our study, prohibitive language in state scope of nursing documents was associated with hospital level protocols for laboratory tests, x-rays, over the counter medication administration, and ECGs. These simple procedures are essential to expediting patient care, and there is specialty training available for emergency nurses to ascertain competency and skill in nurse-initiated protocols (ENA, 2014).

The number of issues raised by the participants in their qualitative comments speaks to the lack of standardization or clarity regarding the use of nurse-initiated protocols. Legislative, regulatory or institutional authority for a hospital to create protocols is not clear. Established pathways for the approval of nurse-initiated protocols that include the input of both medical and nursing staff along with other departments (financial, legal) that can assess the desirability, appropriateness, and efficacy of utilizing nurse-initiated protocols is lacking. Types of tests, medications, treatments that can improve the quality of care delivered to the patient and enhance the patient’s ED experience are undefined by the nurses’ comments. Nurses who made comments in response to the question as to whether they could initiate protocols describe a situation that is inconsistent, unclear, and often raises their concern that they may be practicing beyond their scope of practice as they understand it to be.

Previous research supports the benefits of nurse-initiated protocols to reduce treatment times and increase ED efficiency (Bruse et al., 2015; Considine et al., 2019; Douma et al., 2016; Ho et al., 2018; Retezar et al., 2011; Rivers et al., 2001; Rowe et al., 2011). Several quality metrics depend on timely care initiated immediately upon the patient’s arrival to the ED (Leveille et al., 2020; Sepahvand et al., 2019; McClelland et al., 2012). Previous research also indicates that nurses will violate their own scope of practice if their actions will enhance quality care and enhance efficiency, putting the needs of the patient above their own legal protection (Castner, 2011; Hughes, 2012). Our study has important implications on the need to standardize nurse scope of practice, in relation to initiating emergency care, for both patient care quality and protection of professional nurses who are dedicated to providing the highest quality of care. One key to transforming the health care system to better quality, efficiency, and cost-effectiveness is to consistently allow nurses to practice to the full extent of their education and training by removing regulatory barriers (Committee on the Robert Wood Johnson Foundation Initiative on the Future of Nursing, 2011).

To the best of our knowledge, there has not been a similar survey or policy analysis completed in regards to nurse scope of practice and hospital policy for emergency nursing. The findings of this study underscore that state licensing and regulation are not all well aligned with the nurses’ education and training. Regulatory barriers to timely care leads to inequality and reduces efficiencies within the ED, directly impacting patient outcomes (Lawrence et al., 2007). Elimination of legal and regulatory barriers to the adoption of nurse-initiated protocols would not ensure facilities would adopt the use of nurse-initiated protocols. Physician resistance, inadequate nurse training, cost considerations or other factors could limit use even with consistency in state scope of practice laws favoring the use of nurse-initiated protocols. Nevertheless, state scope of practice laws must be reformed to a national standard that promotes the adoption of evidence based practice nationwide so that RN practice may evolve to meet the changing needs of society and the evolution of the health care delivery system.

## Limitations

The results of this study should be interpreted in light of several limitations. Attempting to reach professionals working in a high-stress, high-demand environment, we created the briefest possible questionnaire for the purpose of this study and did not use an instrument with previous psychometric testing. Secondly, we recruited participants through professional nursing organizations and social media, increasing the potential for response bias and a lack of adequate and unbiased representation across hospital settings. Thirdly, while a very small number, six surveys reported the exact same hospital characteristics as other surveys. This potentially violates the independence of observation assumption of the logistic regression analysis. Finally, it is not possible to transfer or generalize the qualitative comments within this study as the comments came from open-ended survey questions since not every participant answered these questions (a second potential for response bias) and these opinions were not independently verified (as one might in an interview).

## Conclusion

In this study, we explored the relationship of the state-level prohibition of nurse-initiated protocols and hospital application of nurse-initiated protocols relevant to emergency care. We demonstrated a statistically significant relationship between hospital policy and state nurse scope of practice language. Regulatory barriers, such as prohibition of nurse-initiated protocols that do not allow nurses to practice to the full extent of their education and training may limit the quality and efficiency of life-saving emergency care in several states. Additional research is warranted to further elucidate the implications of delayed treatment in the ED that results from prohibitions on nurse-initiated protocols.

## Data Availability

Data not available due to restrictions

